# Day Camp in the Time of COVID-19: What Went Right?

**DOI:** 10.1101/2021.03.11.21253309

**Authors:** Sharon Nachman, Gabrielle Brauner, Aviva Beleck, Andrew S. Handel

## Abstract

**Objective:** To evaluate whether a successful camp experience can be achieved with implementation of COVID-19 education, screening and hygiene protocols, and designated cohorts during the summer of 2020.

**Study Design:** A survey study of summer day camp directors in the metro-New York area was conducted in September, 2020. The survey inquired about camper demographics, COVID-10 related policies, and the number of COVID-19 cases and exposures at each camp.

**Results:** Responses were received from 77% (23/30) of camp directors at the completion of the 2020 summer. There were 8,480 camper children and 3,698 staff across the 23 camps surveyed. A variety of precautions were taken to limit COVID-19 incidence among campers and staff, most often including COVID-19 screening at entry, cohorting campers, maximizing outdoor activities, mandating mask use when indoors, and frequent hand sanitizing. Six staff and one camper tested positive for COVID-19. There was no secondary spread within the staff or campers in any of the camps.

**Conclusion:** Camps successfully stayed open in the summer of 2020. The low level of COVID-19 in the community was critical to the initial success of camp opening. Policies that were consistent and maintained among the camps helped prevent further spread.

## Introduction

Severe Acute Respiratory Syndrome Coronavirus 2 (SARS-CoV-2) emerged in December 2019, quickly leading to a global health crisis. As of August 2020, there have been 20 million cases and over 750,000 deaths internationally^1^. In March 2020, many states in the United States closed schools, nonessential businesses, and public gatherings in an attempt to slow the virus’s spread. In May and June 2020, states began to gradually reopen these businesses and organizations. On June 1, New Jersey’s Governor Murphy announced that summer day camps could open on July 6.^2^ On June 2, New York State’s Governor Cuomo announced that summer day camps would be permitted to open as of June 29.^3^ The United States Centers for Disease Control and Prevention (CDC) published guidelines for safely reopening camps, which included separating campers into non-mixing cohorts, encouraging six-foot social distancing, prioritizing outdoor activities, isolating campers when ill, enforcing mask use, and encouraging proper hand hygiene and sanitation^4^.

Long Island Camps and Private Schools Association (LICAPS) is an organization of twenty-two licensed summer day camps and private schools in Nassau and Suffolk County, New York, typically attended by hundreds of children and staff members^5,6^. Many LICAPS affiliates adopted policies and protocols for reducing COVID-19 cases among campers and staff, and subsequently opened for day camp during the summer of 2020. Investigation of how well these camps performed with regard to identifying COVID-19 cases, preventing virus spread, and adhering to pre-determined camp policies and CDC recommendations will have significant implications for reopening schools and maintaining an environment free of COVID-19.

## Methods

We designed an original survey (Supplement 1) for distribution to the directors of day camps affiliated with LICAPS or located in other metro-New York regions. The survey inquired about general camp demographics, camp policies regarding COVID-19 screening and prevention, and actual illnesses occurring among campers and staff during the 2020 summer. The survey was distributed to all LICAPS camp directors, and other camp directors through three direct email messages over the course of one week in early September, 2020. Of the camps that did not submit a completed survey, specific targeted questions regarding camp size and COVID-19 events were requested and received.

Our study was evaluated by the Stony Brook University Hospital institutional review board and deemed exempt. Study consent was assumed based upon completion of the survey instrument. Results were analyzed using descriptive statistics.

### Analysis

Working with LICAPS allowed us to contact many camps easily. This type of convenience sampling made it possible to reach a large number of camps quickly. Once the surveys were submitted, we were able to download the responses and analyze them for commonalities and outliers.

## Results

### Camp Demographics

23 of 30 camp directors invited to participate completed the survey (77% response rate). Of the camps that did not complete the survey, all responded to specific questions sent to the camp directors requesting limited information. Of those camps, all had an attendance size that fell within the ranges of the group that did respond.

Overall attendance was lower in 2020 than 2019, with a total 2020 attendance of 8,480 camper children and 3,698 staff across the 23 camps who completed the full survey. Attendance in 2019 ranged from 300 to 1,300 campers per camp, compared to 100 to 1,000 campers in 2020 (Figure 1). Compared to 2019, 12 camps had the same number of staff members, while the remaining 11 camps hired fewer staff members. Staff size decreased on average by 83% (range decrease by 525% to an increase by 4%).

**Figure 1.**
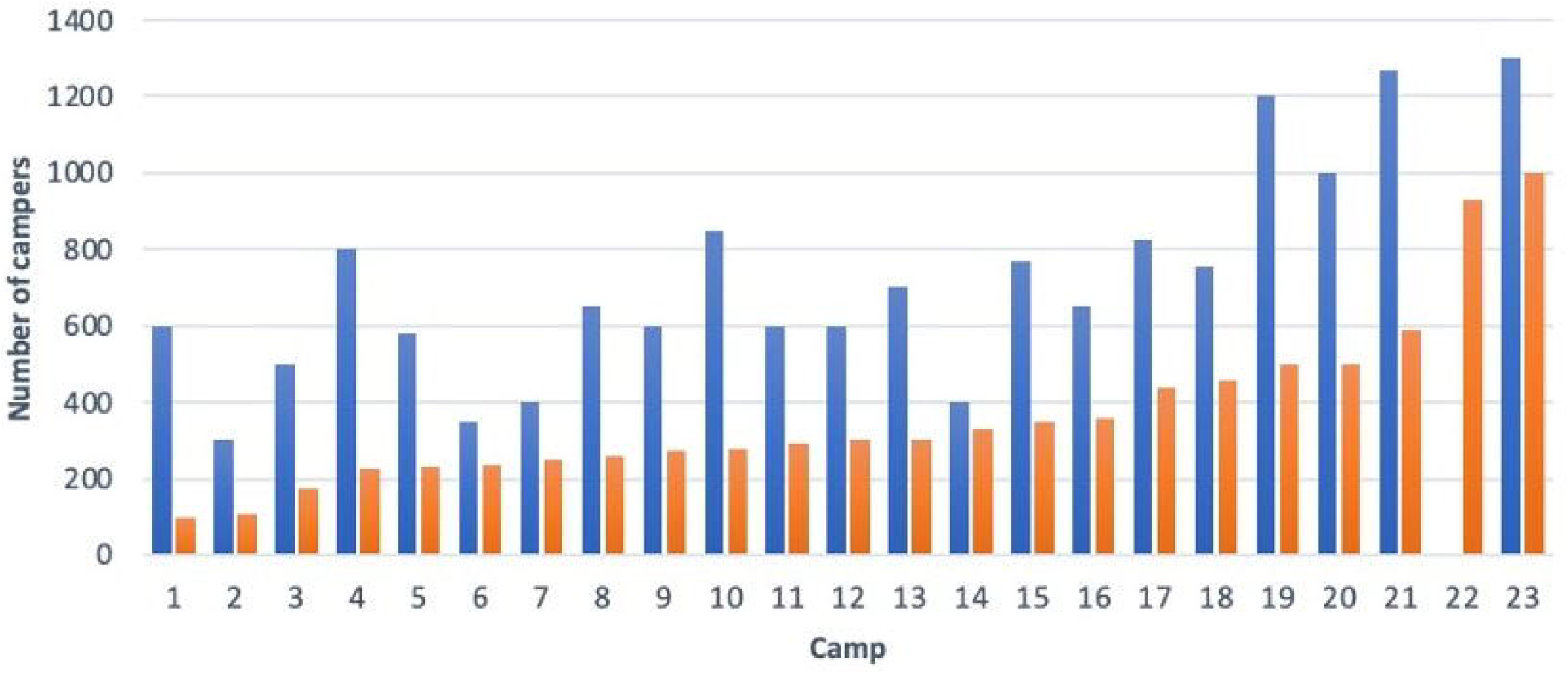
Number of campers in attendance at each surveyed camp in 2019 and 2020

On average, the proportion of campers in each of the designated age cohorts did not vary between 2019 and 2020: 23% of campers in 2020 were in Pre-Kindergarten/Kindergarten, (range 0-55%), 53% were in Elementary School (range 38-55%), and 23% were in Middle School/High School (range 0-58%). All of the camps had similar numbers of males to females per age group.

### Cohorting

Each of the camps surveyed employed closed cohort methodology to limit mixing between large groups of children. Camps maintained an average of 12 campers per cohort (range 3-30). Two camps allowed limited mixing across cohorts. One allowed mixing during drop off and pick up and the other allowed limited age-specific mixing during lunch and swim time. All other camps surveyed did not permit any mixing between the cohorts.

### Facilities

All 23 camps conducted camp outside at least 75% of the time. Regularly used indoor facilities included changing rooms, activity areas, dining areas, health center, and bathrooms. When inclement weather was expected, 16 camps cancelled camp for the day, while others held indoor activities. Camps that cancelled for poor weather conditions missed an average of three days during the summer (range 1-7 days). When inclement weather developed during the day, nine camps held camp inside. There were no field trips in 2020. All camps except one had children change clothing indoors. 90% of the camps had campers eat meals outdoors, with half (10/21) requiring six-foot social distancing while eating.

### Screening

Protocols for pre-camp COVID-19 screening varied. 11 camps required proof of a negative COVID-19 nasopharyngeal PCR from all campers within one week of the start date. Of the 12 camps not requiring a negative COVID-19 PCR, nine required that campers have no COVID-19 symptoms during the 2 weeks prior to starting camp, and one other camp required campers have no contact with SARS-CoV-2 infected individuals for two weeks prior to camp start. The remaining two camps did not require a negative COVID-19 PCR and did not provide details about pre-camp screening protocols.

Daily COVID-19 screening included a mandatory temperature check at all camps. In addition to temperature screening upon arrival onto the campgrounds, 10 camps also required morning home temperature screens, and one camp screened campers’ temperatures on the morning bus. 20/23 (87%) of the camps also asked parents to complete daily symptom screening questionnaires.

### Transportation

Each camp allowed private drop off for the arrival of the campers. In addition, seven of the camps also employed the use of buses, of which five cleaned the buses after every use and two cleaned the buses once daily.

### Masking and Hand Hygiene

78% (18/23) of camps required some form of masking. The majority of camps (78%) required campers to wear a mask whenever inside. Two additional camps required campers to wear a mask when indoors for a prolonged time (i.e. for an activity, but not during change for swim). Only one camp required campers to wear masks all day. All camps except for one (96%) required staff members to wear masks. 65% of camps required staff members wear masks at all times, and 30% only required staff members wear masks when inside. All camps enforcing mask use did provide one when a camper or staff member did not bring one.

All 23 camps supplied hand sanitizer at multiple stations on their campgrounds, as well as soap and water near bathrooms, changing rooms, and the activity areas. Hand sanitizing by campers was required upon arrival to the campgrounds by 70% of camps, before and after activities by 87% of camps, and before eating by 91%. 82% of camps required staff and campers to carry sanitizer with them at all times. All camps reported sanitizing the camp on a nightly basis, though specific cleaning regimens were not reported.

### COVID-19 Symptomatic & Exposed Individuals

All camps required campers or staff members with fever, shortness of breath, and/or cough to leave the campgrounds immediately. Most camps (78% for campers, 70% for staff) required documented clearance from a healthcare provider prior to returning. The majority of camps (83% for campers, 70% for staff) also required a two-day symptom-free period prior to returning. Only one camp required a negative COVID-19 assay for all sick campers prior to returning to camp. Regarding staff, one camp required all symptomatic staff members quarantine at home for two weeks prior to returning, and two camps required staff members demonstrate a negative COVID-19 test before returning. 11 camps required a note from the staff member’s doctor and the staff member to be asymptomatic for 2 days before returning to camp.

The camps all developed protocols regarding return to camp for COVID-19 infected individuals. Most camps required infected individuals have a 14-day home isolation period after testing positive (82% for campers, 78% for staff) and a documented negative COVID-19 PCR (86% for campers; 78% for staff) prior to returning to camp. 74% of camps also required written clearance to return to camp from the camper’s healthcare provider.

### Illness events

Excluding symptoms due to laboratory-confirmed COVID-19, only 57/12,178 individuals developed an illness during a camp day. Daily, each camp had an additional 1-2 campers or staff out of attendance due to feeling ill. None of these resulted in a new COVID-19 diagnosis, and all required either written clearance from a healthcare provider, a COVID-19 test, or both prior to re-entry. Overall, the most common reasons campers and staff were sent home included fever (13 campers and 19 staff members) and vomiting or diarrhea (11 campers and 14 staff). Symptoms concerning for COVID-19 accounted for sending 10 campers and 5 staff members home. COVID-19 exposures accounted for 4 campers and 3 staff being sent home. None of these individuals were COVID-19 positive.

### Documented COVID-19 cases

COVID-19 cases occurred in two camps. The first camp documented 1 camper and 5 staff members (with a common exposure outside of camp) with positive COVID-19 PCR testing, with an infection rate of 1.4%. The individual camper was ill on day 2 of camp, suggesting a prior outside exposure as well. One staff member, several days into the season, was diagnosed with COVID-19 at the second camp, giving an infection rate of 0.08%. Both infection rates were well below the local infection rate at that time, which were under 3% for the county where the first camp is located^7^ and under 7% for where the second camp is located.^8^ Of the camps that did not complete the survey, one camp also documented several cases of COVID-19. These individuals (4 campers and 8 staff members) tested positive around the 3^rd^ week of camp. They were sent home to be isolated, the camp was closed for 1 week, and no further campers or staff were infected.

## Discussion

Our understanding of COVID-19 transmission has changed countless times since the pandemic was first recognized. Among the many unresolved questions is the extent to which minimally symptomatic or entirely asymptomatic children contribute to the virus’ propagation. As the vast majority of infected children exhibit few signs of illness^9^, a clear challenge exists in safely restarting schools and camps while minimizing inadvertent spread of the virus to other children, teachers, and the greater community. Our study found that, in a region with relatively well-controlled COVID-19 spread, and with close attention to prevention policies, new infections were uncommon among day camp attendees and staff. Further, the single outbreak that did occur was quickly halted, with only a handful of individuals infected, all from outside of the camp activities. A similar outcome, lack of spread among campers and staff, occurred in the camp with little in-depth information. Although not a controlled study, the tactics used by these camps may assist other childhood institutions in designing protocols for safely reopening.

A review of camp policies showed a high degree of similarity. This was due to industry-shared protocols. Although some policies may have differed between camps, these were limited. These included across all camps: decreasing the number of campers (due to a self-selective process of which families wanted to participate in camp), requiring staff members to wear masks when inside, isolating any person with any illness or COVID-19 symptoms or exposure. Other common policies included conducting camps in cohorts, holding most camp activities outside, daily temperature screening, providing ample hand sanitizer, and cleaning the camp nightly. The degree to which each of these strategies limited COVID-19 episodes in the camp cannot be determined.

Pivotal to the success of camp reopening was the low rate of infection in the community. Throughout the summer (starting in June), the percent positivity was under 3% for Suffolk County^7^, Nassau County^7^, and Somerset County^8^, and under 2% for New York County ^7,^ Westchester County^8^, and Rockland County^7^. Underscoring this point, the camp with one diagnosed COVID-19 case occurred in Burlington County, which had a much higher infection rate of 7%.^8^

Contact tracing and prevalence studies have suggested that those younger than 10-14 years old might be less susceptible to COVID-19 infection than those 20 years of age and older. There is also evidence that adolescents and children might contribute a smaller role in transmission of COVID-19 than people of other ages.^10^ These would, in part, help explain the low rates of infection seem among day camps.

As a side product of masking of staff, screening of campers, and good hand hygiene, very low rates of other illnesses were seen as well. Most summers, cases of Coxsackievirus, Streptococcal pharyngitis, and acute viral gastroenteritis are common (personal communication Mark Transport, LICAPS President). This past summer, however, there were no cases of Coxsackievirus or Streptococcal pharyngitis among the 8,480 camper children and 3,698 staff members across the 23 camps surveyed.

What was difficult to quantify is the level of loyalty to the camp. Parents, staff, and campers all appreciated that what they did in and out of camp hours directly influenced the ability of the camp to succeed. Often campers and staff had a long-standing relationship with the individual camps, and a sense of community with that camp. By explicit and implicit discussion, they all understood that attendance in camp was a privilege and that disregard of social distancing, mask wearing, and hand hygiene rules both inside and out of camp put continued attendance and success at risk.

Critical to success was ongoing good communication between staff, parents, and camp directors. Frequent contact between camp directors, staff, and parents permitted ongoing discussion and reinforcement of the camp’s COVID-19 rules. Sleep away camps, which were prohibited from opening in New York State but allowed to open in twenty-five other states,^11^ have also provided valuable insights into reopening during the pandemic. Notably, numerous sleep away camps throughout the United States were closed after opening due to COVID-19 outbreaks, despite following state specific guidelines^11,12,13,14^. Good communication was not always the norm for these camps as compared to the summer day camps in our survey. Typical examples of these poor communications included lack of notification to families when a camper tested positive or holding large group sessions to notify campers that a COVID-19 outbreak had occurred, risking further spread.^12^ Additionally, one large camp of 7,000 campers seemingly did not adhere to its own social distancing policies.^13^ Just as with camps, reopening schools are now exploring optimal strategies for minimizing COVID-19 cases, and they may look to camp successes for guidance. One crucial difference is the general reliance of schools on indoor activities. Schools may benefit from strategies recommended by the CDC and employed by the camps, namely enforcing proper hand hygiene, mask use, social distancing, and student cohorting, facilitating frequent COVID-19 testing, and quickly isolating COVID-19 infected and exposed individuals. Frequent school-wide sanitizing and closures following a single case has been implemented since school reopening, though these practices are not evidence-based at this time.

## Conclusions

When a decision was made to allow day camps to reopen in New York in summer 2020, many parents feared putting their children at risk by sending them to camp. We report here data from 23 camps which responded to a survey regarding camp activates. The majority of the summer camps surveyed had no positive COVID-19 cases identified. Two summer camps who completed the full survey, and one additional camp who completed a partial survey had positive cases. There was no spread within the camps due to fast recognition and action. These camps were successful because of a low level of virus in the community, effective screening practices, emphasizing outdoor activities, and proper hygiene and masking. With these examples of successful summer camp experiences, we are hopeful that safe school reopening and limiting COVID-19 spread within the school and the community at large can be maintained.

## Data Availability

Study data is maintained by the study team and available upon request.

## Acknowledgements

The following camps participated in the survey. We thank all of their staff and leadership for providing us with the data described in the manuscript. In alphabetical order:

Breezemont Day Camp (Gordon Josey), Brookhaven Country Day Camp (Michael Pollack), Camp Jacobson (Paul Isserles), Camp Nabby (Rita C. Bertino), Crestwood Day Camp (Mark Transport and Mark Hemmerdinger), Deerkill Day Camp (Todd Rothman), Driftwood Day Camp (Mike Wagenberg and Ron Kuznetz), Elmwood Day Camp (Gregg Licht), Hidden Pond Day Camp (Matt Pagliari), Ivy League Day Camp (Meredith Stern), Kenwal Day Camp (Howard Feinstein), Liberty Lake Day Camp (Andy Pritikin), Maplewood School & Summer Program (Sheri Holden), Mohawk Day Camp (Adam Wallach), North Shore Day Camp (Joni Iacono), Oak Crest Day Camp (Jonathon Gold), Park Shore Country Day Camp (Robert Budah), Pierce Day Camp (Will and Courtney Pierce), Rolling River Day Camp (Marissa Allaben and Mark Goodman), Shibley Day Camp (Heath Levine), Summer Trails Day Camp (Jamie Sirkin), The Nature Place Day Camp (Scott Dunn), Woodmont Day Camp (Sam Borek)

## Abbreviations

SARS-CoV-2: Severe Acute Respiratory Syndrome Coronavirus 2
CDC: Centers for Disease Control and Prevention
LICAPS: Long Island Camps and Private Schools Association

## Supplement Legend

Supplement 1: Complete survey instrument

## Notes

**Potential Conflicts of Interest:** None

### Competing Interest Statement

The authors have declared no competing interest.

### Funding Statement

This work was completed without any specific funding source.

### Author Declarations

Our study was evaluated by the Stony Brook University Hospital institutional review board and deemed exempt.

