## Supplement 1 for "Day Camp in the Time of COVID-19: What Went Right?"

**Basic Camp Demographics:**

Name of the camp

State that camp is in

- New York
- New Jersey

County that camp is in

Name of person filling this out

What date did camp begin this year?

What date will camp end this year?

How many hours is camp in session?

- 1-3
- 4-6
- 7-9

**This year:**

Number of campers this year

Number of campers in preschool-kindergarten

Number of campers in elementary school

Number of campers in middle school-high school

Number of staff members this year

Number of staff members younger than 18

Number of staff members 18-25

Number of staff members 25+

Number of staff members returning this year

◯ Same number of returning staff as prior years

◯ Fewer returning staff than prior years

**Prior years:**

Number of campers in prior years

Number of campers in preschool-kindergarten in prior years

Number of campers in elementary school in prior years

Number of campers in middle school-high school in prior years

Number of staff members in prior years

Number of staff members younger than 18 in prior years

Number of staff members 18-25 in prior years

Number of staff members 25+ in prior years

### Camp Policies

Prior to camp reopening, negative Covid-19 test was requried

- ◯ Yes
- ◯ No

If a negative test was not required, what did you require? Check all that apply

- ☐ No symptoms within 2 weeks of camp
- ☐ Other
- ☐ Quarantine for 2 weeks prior to coming to camp

When was negative Covid-19 testing required

- ◯ Within 1 week of the start of camp
- ◯ Within 2 weeks of the start of camp

Camp is conducted in cohorts

- ◯ Yes
- ◯ No

What is the average number of children in each cohort

What is the smallest cohort

What is the largest cohort

Is there mixing of cohorts during drop off and pick up

- Yes
- No

Is there mixing of cohorts during lunch

- Yes
- No

Is there mixing of cohorts during swim

- Yes
- No

No mixing of cohorts is allowed

- Yes
- No

Camp is conducted outside at least 75% of the time

- Yes
- No

When there is expected inclement weather, camp is cancelled

- Yes
- No

When there is expected inclement weather, camp is conducted inside

- Yes
- No

How many days of expected inclement weather were there this summer

Please check which indoor facilities were used. Check all that apply

- Changing rooms
- Dining areas
- Activity areas
- Other

**Screening**

Temperatures are screened daily

- Yes
- No

Where? Check all that apply

- Home
- Other
- Camp

Screening questions are asked of parents daily

- Yes
- No

**Transportation**

How did the campers get to camp? Check all that apply

- By bus
- Other
- Private drop off

How often are buses cleaned?

- After every use

- Only once daily
- They are not cleaned

**Masking**

Staff are required to wear a mask: Check all that apply

- All day
- When inside only
- When outside only when within 6 feet of another person
- Not at all

What type of mask is allowed? Check all that apply

- Any face covering
- Gaiter mask
- Surgical mask
- Other
- Cloth mask

Campers are required to wear a mask: Check all that apply

- All day
- When inside at all, even for short periods of time
- When outside only when within 6 feet of another person
- Not at all
- When inside for extended periods of time

What type of mask is allowed? Check all that apply

- Any face covering
- Gaiter mask
- Surgical mask
- Other
- Cloth mask

Campers are supplied with masks if they show up without one

- Yes
- No

What type of mask is supplied? Check all that apply

Surgical mask
Gaiter mask

- [ ]
- [ ]
- [ ] Cloth mask
- [ ] Other ____

Campers are sent home if they show up without a mask

- ( ) Yes
- ( ) No

Staff members are supplied with masks if they show up without one

- ( ) Yes
- ( ) No

What type of mask is supplied? Check all that apply

- [ ] Surgical mask
- [ ] Gaiter mask
- [ ] Cloth mask
- [ ] Other ____

Staff members are sent home if they show up without a mask

- ( ) Yes
- ( ) No

### Sanitation

How is hand cleanliness maintained, please check all that apply

- [ ] Hand sanitizer stations at camp
- [ ] Soap and water are available

Where were the stations? Check all that apply

- [ ] By bathrooms
- [ ] By changing rooms
- [ ] By activity areas
- [ ] Other ____
- [ ] By dining areas

Where are the sinks located? Check all that apply

- ☐ By bathrooms
- ☐ By changing rooms
- ☐ By activity areas
- ☐ Other ________
- ☐ By dining areas

When are hands washed? Check all that apply

- ☐ On arrival
- ☐ After eating
- ☐ Before activity
- ☐ After bathroom
- ☐ After activity
- ☐ Staff and campers carry sanitizer with them at all times
- ☐ Before eating

The camp is sanitized every night

- ○ Yes
- ○ No

**Activities**

There are field trips

- ○ Yes
- ○ No

Is there a lake/river

- ○ Yes
- ○ No

Is there a pool

- ○ Yes
- ○ No

Do children change for swim inside

- ◯ Yes
- ◯ No
- ◯ N/A

Are campers required to wear a mask while changing for swim

- ◯ Yes
- ◯ No

### Food and Drink

Please check all that apply

- ☐ Campers bring lunch from home
- ☐ Lunch is provided for the campers
- ☐ Lunch is served in a cafeteria
- ☐ Campers serve themselves the provided lunch
- ☐ Lunch is eaten outside

Are campers socially distant when they are eating lunch/snacks

- ◯ Yes
- ◯ No

Please check all that apply

- ☐ Campers bring their own water
- ☐ Water is provided for the campers
- ☐ Water bottles are provided for the campers
- ☐ There is a common water cooler for campers to use

### Illness Policies in Camp

Camp policy if a camper has a fever upon arrival or during the day (and did not have a fever before coming to camp)
Please check all that apply

- [ ] Camper is sent home right away
- [ ] Camper is not isolated and is kept at camp
- [ ] Camper is isolated, but kept at camp
- [ ] Other ______

Camp policy if a camper tests positive for Covid-19 before they are allowed back to camp, they need:
Please check all that apply

- [ ] COVID negative test
- [ ] Note from pediatrician
- [ ] Camper has to quarantine for 2 weeks
- [ ] Other ______

Camp policy if a camper is exposed to Covid-19
Please check all that apply

- [ ] Camper is sent home right away
- [ ] Camper is not isolated and is kept at camp
- [ ] Camper is isolated, but kept at camp
- [ ] Other ______

Camp policy for a camper with COVID symptoms (such as fever, cough, shortness of breath) but is COVID negative. Before they are allowed to return to camp, they need:
Please check all that apply

- [ ] Note from pediatrician
- [ ] Camper has to be without symptoms for 2 days before returning
- [ ] Camper has to quarantine for 2 weeks
- [ ] Other ______

Camp policy if a staff member has a fever upon arrival or during the day (and did not have a fever before coming to camp)
Please check all that apply

- [ ] Staff member is sent home right away
- [ ] Staff member is not isolated and is kept at camp
- [ ] Staff member is isolated, but kept at camp
- [ ] Other ______

Camp policy if a staff member tests positive for Covid-19, before they are allowed back to camp, they need:
Please check all that apply

- [ ] COVID negative test
- [ ] Note from doctor
- [ ] Staff member has to quarantine for 2 weeks
- [ ] Other ______

Camp policy if a staff member is exposed to Covid-19:
Please check all that apply

- [ ] Staff member is sent home right away
- [ ] Staff member is not isolated and is kept at camp
- [ ] Staff member is isolated, but kept at camp
- [ ] Other

Camp policy if a staff member has COVID symptoms (such as fever, cough, shortness of breath) but is COVID negative. Before they are allowed to return to camp, they need:
Please check all that apply

- [ ] Note from doctor
- [ ] Staff member has to be without symptoms for 2 days before returning
- [ ] Staff member has to quarantine for 2 weeks
- [ ] Other

Did you have a COVID coordinator: a staff member dedicated to overseeing COVID related things?

- ○ Yes
- ○ No

**Illness Events in Camp**

How many campers were sent home from camp this summer

- ○ 1-5
- ○ 6-10
- ○ 11-15
- ○ 15+

Reasons for being sent home, please check all that apply

- [ ] Fever
- [ ] GI illness
- [ ] Covid-19 EXPOSURE
- [ ] Covid-19 SYMPTOMS (shortness of breath, cough, sore throat, runny nose, body aches, loss of smell, fatigue, headache)

Please select which answer choice best applies: How did your cases of coxsackie virus, diarrheal illnesses, and colds this summer compare to last summer?

- ○ Significantly more than last summer
- ○ More than last summer
- ○ Same as last summer

- ◯ Less than last summer
- ◯ Significantly less than last summer

How many staff were sent home from camp this summer

- ◯ 1-5
- ◯ 6-10
- ◯ 11-15
- ◯ 15+

Reasons for being sent home, please check all that apply

- ☐ Fever
- ☐ GI illness
- ☐ Covid-19 EXPOSURE
- ☐ Covid-19 SYMPTOMS (shortness of breath, cough, sore throat, runny nose, body aches, loss of smell, fatigue, headache)

Were there any Covid-19 infections identified in campers this summer

- ◯ Yes
- ◯ No

How many

________________

How many cohorts were involved

________________

Were there any Covid-19 infections identified in staff this summer

- ◯ Yes
- ◯ No

How many
